# Therapeutic inertia and blood pressure control status among adult hypertensive patients in Ethiopia: Prospective observational study

**DOI:** 10.1101/2025.09.18.25336077

**Authors:** Musse Gelebo Garo, Tsegaye Melaku

## Abstract

Hypertension is a leading cause of death and disability worldwide, including in developing countries like Ethiopia. The causes may include therapeutic inertia, patient-related risk factors, or medication nonadherence. Therefore, this study aimed to assess the burden of therapeutic inertia and the blood pressure control rate among adult hypertensive patients on follow-up at Jimma Medical Center. A prospective observational study was conducted among adult hypertensive patients who had regular follow-ups at the Ambulatory Cardiac Clinic of Jimma Medical Center from September 22 to December 22, 2022. Patients’ specific data were collected using a structured data collection tool and through face-to-face interviews. Blood pressure control status was evaluated following the Eighth Joint National Committee guideline recommendations. Data were analyzed with SPSS version 27.0. Factors associated with blood pressure control were identified using binary and multivariate logistic regression analyses. Among 360 participants, 208 (57.8%) were male with a mean age of 54 ± 13.523 years. The blood pressure control rate was 36.9%, and the medication adherence rate was 47.2%. The three-month mean therapeutic inertia score was 0.32 ± 0.51, with over half (56.9%) of participants having a higher therapeutic inertia score than the mean. In multivariable logistic regression analysis, not reducing salt intake (*P* = 0.003), medication nonadherence (*P* = 0.009), therapeutic inertia scores during the first, second, and third visits (*P* = 0.001, 0.013, and 0.041, respectively), and a higher overall therapeutic inertia score (*P* = 0.013) were significantly associated with uncontrolled blood pressure. The rate of blood pressure control was low. Non-salt reduction, medication nonadherence, and therapeutic inertia were independent predictors of blood pressure control. Therefore, clinicians should adhere to guidelines to reduce therapeutic inertia and provide health education on salt consumption and medication adherence.

## Introduction

Hypertension remains a major factor contributing to mortality worldwide. It imposes a significant economic burden globally, responsible for nearly 1 in 5 deaths worldwide and 1 in 11 in low-income countries. Poorly managed hypertension is a leading cause of hypertension-related complications, including cardiovascular disease (CVD) and death from chronic renal disease. The impact of high blood pressure (BP) varies by region, age, and gender (1–3). Inadequately treated hypertension accounts for over 50% of heart disorders and strokes, resulting in 10.8 million deaths (19.2% of all deaths) in 2019 globally (4, 5).

Much of the hypertension burden has shifted from high- and middle-income regions to lower-income areas, creating a public health crisis in sub-Saharan Africa (SSA) (1, 6). Hypertension affects millions of people in SSA and, if left unaddressed, is a leading cause of cardiovascular disorders and stroke (7). Despite the availability of cost-effective lifestyle changes and medical treatments that can control hypertension and reduce mortality and disability, the African continent still has a relatively high prevalence of the condition, with poor rates of detection, treatment, and BP control (8). In eastern SSA, the average prevalence of hypertension was 20.95%. Additionally, in this region, the rates of BP control and medication adherence were low at 11.5% and 60%, respectively, mainly due to failure to follow prescribed medications, lack of awareness, unavailability of medicines and medical support, weak healthcare providers, and the absence of evidence-based guidelines (9).

Increased health risks, such as stroke, heart failure (HF), and kidney failure, are expected to occur in Ethiopia over the coming years, as the prevalence, distribution, and mortality rates are all alarmingly high (10). Studies conducted in Ethiopia show that the overall rate of hypertension and uncontrolled hypertension is 20.63% and 48%, respectively. This significant public health issue contributes to the rising incidence of cardiovascular and cerebrovascular diseases (11, 12). In Ethiopia, the study indicates that hypertension was responsible for 5.6% of all deaths and was among the top five causes of cardiovascular disease-related deaths in 2017 (13).

Originally, clinical inertia described gaps in practice; now, therapeutic inertia (TI) is often used interchangeably with clinical inertia. TI refers to healthcare providers’ inability to initiate or escalate treatment as recommended by guidelines (14). This issue is increasingly recognized as a major reason for clinicians’ failure to effectively manage hypertension, leading to a higher risk of cardiovascular events (15). It prevents therapy intensification and significantly contributes to low hypertension control rates (16). Antihypertensive treatment typically involves gradual up-titration based on BP readings. However, reaching the target is difficult due to TI (17). Numerous studies worldwide have shown that TI is common and hampers BP control. In Africa, some studies found that only 10% of patients with uncontrolled BP had treatment changes made (18–22). Research also indicates that physician-related barriers, such as TI, have not been thoroughly studied, despite being critical for achieving BP control. The prevalence of TI in such cases ranges from 80% to 90% (23).

Controlling and preventing high BP is the main approach to prevent and manage non-communicable diseases, serving as an example of addressing other non-communicable disease risks (24). Enhancing medication adherence may significantly impact our community’s well-being more than discovering new treatments (25). However, adherence to hypertension medication remains low and is linked to an increased risk of future cardiovascular events, especially among young patients (26). Numerous studies have shown that the core issue is a lack of sustained effort and poor daily adherence to recommended therapies (27). The survey in Eastern SSA reported that poor BP control and adherence were due to factors such as low awareness, limited access to medications and healthcare services, clinician reluctance to increase medication doses, lack of understanding of evidence-based guidelines, weak governance, and certain health habits related to rules (9). A systematic review and meta-analysis conducted in Ethiopia found that around 65.41% of patients with high BP adhered to their medications (28). Another systematic review and meta-analysis in Ethiopia indicated that 45% of patients did not adhere to antihypertensive drugs, with a larger proportion (83.7%) having suboptimal BP control and not adhering to treatment (29).

Studies found that several risk factors, such as a family history of hypertension, older age, urban residence, lower educational coverage, diabetes mellitus, BMI, alcohol consumption, physical inactivity, employment status, coffee use, unhealthy diet, khat chewing, and central obesity, are associated with poor hypertension control in Ethiopia (11, 12, 30–33). A study conducted in Kenya showed that uncontrolled hypertension was linked to BMI, alcohol, waist-hip ratio, and tobacco use (34). Another study in Ethiopia identified lower educational coverage, age ≥40, family history of hypertension, urban residence, diabetes mellitus (DM), alcohol consumption, central obesity, and BMI ≥ 25 as risk factors for suboptimal BP control (12).

Several studies worldwide have investigated the effect of TI on BP control (16, 17, 21, 35, 36). However, no data are available regarding the impact of TI on BP control status in Ethiopia, and its association has not been studied there. Additionally, a cross-sectional design has limited most studies on BP control. Therefore, this prospective observational study assessed the burden of TI and BP control among adult hypertension patients on follow-up at the Ambulatory Cardiac Clinic of Jimma Medical Center (JMC).

## Methodology

### Study setting and period

The study was conducted over three months, involving adult hypertension patients who attended routine follow-up appointments monthly at the Ambulatory Cardiac Clinic of JMC from September 22 to December 22, 2022. The hospital is 352 kilometers southwest of Addis Ababa, Ethiopia’s capital. It is located in the Jimma Zone, a teaching and specialized hospital with 660 beds. The hospital treats about 15,000 inpatients, handles 11,000 emergency cases, and conducts 4,500 deliveries annually. Currently, the Ambulatory Cardiac Clinic at JMC provides follow-up care and antihypertensive medication services to around 2,527 ambulatory hypertensive patients (37).

### Study design

A hospital-based prospective observational study was employed.

### Source and study population

The source population for this study included all hypertensive patients undergoing treatment follow-up at the Ambulatory Cardiac Clinic of JMC during the study period. The study population comprised all adult patients with hypertension aged 18 years or older with uncontrolled BP who had at least two clinic visits before the study period, were physically capable and willing to participate, were on at least one antihypertensive medication or combination therapy, and had a clinic visit at the Ambulatory Cardiac Clinic of JMC every month during the study period. Critically ill patients unable to respond, those with mental illness or psychiatric conditions, pregnant women, and individuals with incomplete medical records—such as demographics or BP data—were excluded from this study.

### Sample size and sampling technique

The study recently conducted at JMC reported that the BP control rate was 42.8% (33). Therefore, using the Cochran single proportion formula, the minimum sample size (n) required for this study, based on a standard normal distribution (Z = 1.96) with a 95% confidence interval (CI) and a 0.05 margin of error (E), was calculated to be 376. Since the source population was less than 10,000, the required sample size was adjusted to 327.3. After accounting for a 10% non-response rate, the final sample size was 360. A systematic sampling method was used to select participants from the total patient charts after determining the interval (K). The K-interval was calculated by dividing the total number of charts by the final sample size. The first participant was chosen randomly from a pool of 1 to 7 using a lottery system. The remaining participants were then selected at every kth interval (7) until the target sample size was achieved.

### Data collection tool and procedures

The questionnaire and data abstraction forms were created by reviewing various types of literature (32, 33, 38). To ensure the validity of the data collection tools, a structured questionnaire was developed, translated into local languages spoken by native speakers (Afan Oromo and Amharic), and then back-translated into English. The 9-item Hill-Bone Medication Adherence Scale (HB-MAS) was used to assess participants’ medication adherence. The 9-item HB-MAS does not provide a cutoff point to distinguish adherent from non-adherent patients (39). In this study, participants who reported ‘none of the time’ or ‘some of the time’ were considered to have good medication adherence, while those who reported ‘most of the time’ or ‘all of the time’ were regarded as having poor adherence (40). The Eighth Joint National Committee (8^th^ JNC) guideline recommendations were used to evaluate BP control status. Before starting the study, a pretest of the data collection form was conducted on 5% of the sample. Primary data, such as sociodemographic and lifestyle factors, were collected through face-to-face patient interviews. Additionally, data were obtained directly from clinicians through face-to-face interviews and from patients’ charts regarding TI during each visit. During each visit, prescribed medications, BP readings, dose adjustments, added drug classes, and comorbidities were retrieved from patients’ charts. The attending physician measured initial and follow-up BPs at JMC using an approved method with a validated mercury sphygmomanometer and a cuff appropriately sized for arm circumference, sitting at heart level, after a minimum of 5 minutes of rest. In this study, patients who met the inclusion criteria were recruited from August 17 to September 17, 022, only during the first month of data collection. Therefore, BP levels and antihypertensive medication regimens recorded at this initial appointment served as baseline data. Only those recruited in the first month were monitored monthly for the remaining three months.

### Data processing and analysis

The collected data were reviewed for completeness and consistency. Afterwards, they were edited, cleaned, and entered into the Statistical Package for the Social Sciences (SPSS) version 27.0. The data were displayed and summarized using tables and graphs. Categorical variables showed frequencies and percentages. Continuous variables were reported as mean ± standard deviation (SD). Controlled BP is defined as systolic blood pressure (SBP) < 140 mmHg and diastolic blood pressure (DBP) < 90 mmHg for individuals under 60 years old, and as SBP < 150 mmHg and DBP < 90 mmHg for individuals aged 60 years or older, based on average blood pressure readings. SBP < 140 mmHg and DBP < 90 mmHg for individuals aged 18 years or older with chronic kidney disease (CKD), with or without diabetes, are considered controlled BP. BP was calculated using the average of three measurements and categorized as controlled or uncontrolled according to the 8^th^ JNC guideline recommendations (41). The primary outcome of the study was the rate of BP control and secondary outcomes included the rate of TI and medication adherence rate.

The three-month TI Score for each patient was calculated as follows:

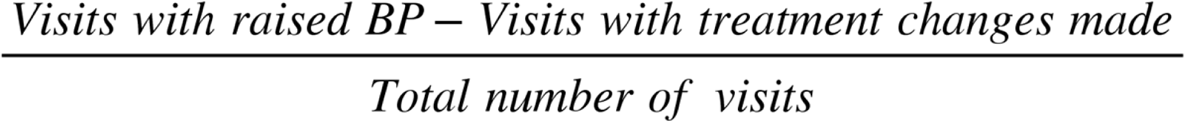

Scoring ranges from -1 to +1, with higher numbers indicating higher TI (21). The logistic regression model assessed the relationship between the dependent and independent variables. Variables with a P-value less than 0.25 in the binary logistic regression analysis were included in the multivariate logistic regression to identify key independent factors affecting BP control. Factors with a P-value below 0.05 were considered statistically significant in the multivariate analysis.

### Ethics approval and consent from participants

Before collecting data, ethical approval of the research protocol was obtained from the Ethical Review Board of the Institute of Health at Jimma University (Ref. No. JUIHIRB/52/22). The hospital also granted permission to conduct the study. After thoroughly explaining the study’s objectives, selection procedures, and confidentiality measures, only participants who met the inclusion criteria were verbally asked if they wished to participate. An independent data-collecting supervisor served as a witness to ensure that participants’ decisions were made freely and with complete understanding. Written informed consent was waived because the study did not involve invasive procedures or patient harm, as the Ethics Review Board Committee of the Institute of Health at Jimma University approved. All interviews were conducted privately, and to protect participants’ confidentiality, their names were not included on the questionnaires. Participants were also informed of their right to refuse to answer some or all questions.

## Result

### Socio-demographic characteristics of participants

As depicted in Fig 1, three hundred ninety-seven hypertensive participants met the inclusion criteria and were invited to participate; ultimately, 360 participants were included in the final analyses, resulting in a response rate of 90.7%.

**Fig 1.**
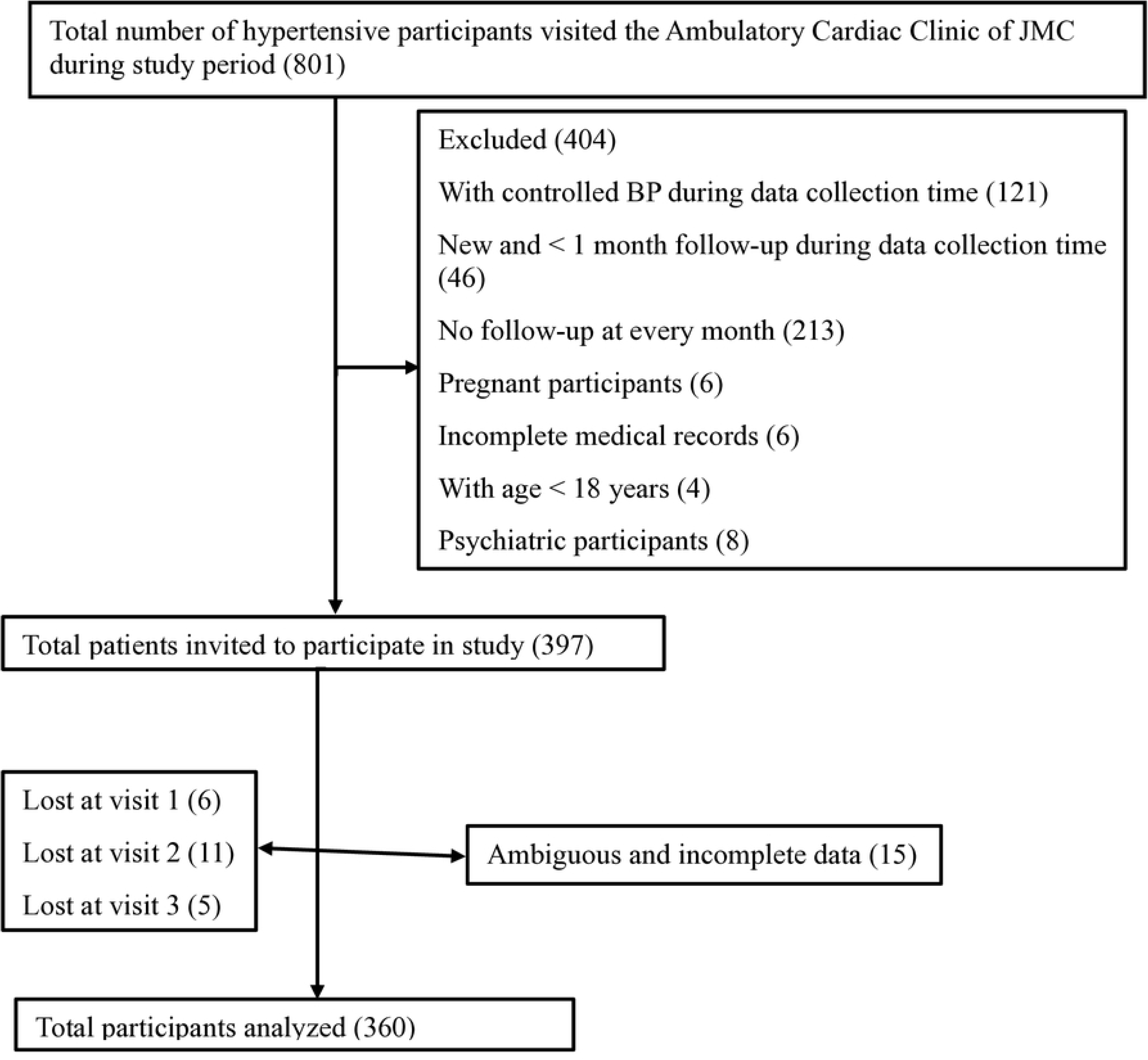
Hypertension participant selection flowchart at JMC from September 22 to December 22, 2022.

As shown in Table 1, 208 (57.8%) participants were male. The mean SD age of participants was 54 ± 13.523 years, with a minimum age of 18 years and a maximum of 85 years. Two hundred ninety-one participants (80.8%) were married, and the majority, 217 (60.3%), resided in an urban area. Among participants, 133 (36.9%) had no formal education, 87 (24.2%) were jobless, and 142 (39.4%) were without a well-defined monthly income. The mean SD BMI of participants was 22.955 ± 2.755 kg/m², with most participants, 290 (80.6%), having normal body weight, and 283 (78.6%) living with their families.

**Table 1.** Socio-demographic characteristics of hypertensive participants at JMC from September 22 to December 22, 2022.

| Variables | Characteristics | Frequency | Percentage |
| --- | --- | --- | --- |
| **Age in years** | Mean SD | 54±13.523 |  |
|  | 18-60 | 195 | 54.2 |
|  | ≥ 60 | 165 | 45.8 |
| **Sex** | Male | 208 | 57.8 |
|  | Female | 152 | 42.2 |
| **BMI** | Mean SD | 22.955±2.755 |  |
|  | 18.5-24.9 | 290 | 80.6 |
|  | 25-29.9 | 58 | 16.1 |
|  | ≥ 30 | 12 | 3.3 |
| **Marital status** | Married | 291 | 80.8 |
|  | Widowed | 28 | 7.8 |
|  | Divorced | 23 | 6.4 |
|  | Single | 18 | 5.0 |
| **Place of residence** | Urban | 217 | 60.3 |
|  | Rural | 143 | 39.7 |
| **Educational status** | No formal education | 133 | 36.9 |
|  | Primary (1–8 grade) | 93 | 25.8 |
|  | Secondary (9–12 grade) | 75 | 20.8 |
|  | Above secondary (diploma and above) | 59 | 16.4 |
| **Current occupation** | Jobless | 87 | 24.2 |
|  | Merchant | 78 | 21.7 |
|  | Employed (Civil servant) | 60 | 16.7 |
|  | Farmer | 45 | 12.5 |
|  | Housewife | 36 | 10.0 |
|  | Retired | 20 | 5.6 |
|  | Others* | 34 | 9.4 |
| **Monthly income in Ethiopian Birr** | Without a well-defined monthly income | 142 | 39.4 |
|  | > 3000 | 105 | 29.2 |
|  | 2001-3000 | 57 | 15.8 |
|  | 1001- 2000 | 30 | 8.3 |
|  | < 1000 | 26 | 7.2 |
| **Living status** | Live with family | 283 | 78.6 |
|  | Live alone | 57 | 15.8 |
|  | Others** | 20 | 5.6 |

Standard Deviation: SD; Body Mass Index: BMI.
Others\*: Drivers, daily laborers, non-governmental organizations, students, and private work.
Others\*\*: Live with friends and relatives, or be in prison.

### Lifestyle, clinical characteristics, and medication adherence of participants

As depicted in Table 2, 189 (52.5%) participants have not reduced the amount of salt in their diet. Most participants, 326 (90.6%), were physically inactive. One hundred thirty-five (37.5%) participants were khat chewers, 14 (3.9%) were current cigarette smokers, 90 (25.0%) drank coffee all the time, and 25 (6.9%) were traditional drug users. Out of the total study participants, 260 (72.2%) had comorbidities; 72 (20.0%) of them had DM, followed by 33 (9.2%) with CKD. Seventy-two (20.0%) participants had other comorbidities like anemia, HF with CKD, peripheral neuropathy, chronic obstructive pulmonary disease, asthma, hypertensive heart disease, arthritis, nephrotic syndrome, dyspepsia, DM with coronary artery disease, gastroesophageal reflux disease, CKD with a history of myocardial infarction, thyrotoxicosis, peptic ulcer disease, and human immunodeficiency virus-acquired immune deficiency syndrome. One-third of the participants (33.3%) had been on antihypertensive therapy for between 1 and 5 years. The baseline means (SD) for SBP and DBP of participants were 148.93 ± 14.47 and 93.95 ± 10.00, respectively, and most of them, 154 (42.8%), were on two-drug combinations. The means (SD) of SBP and DBP for participants during visit one were 146.32±13.36 and 91.23±8.47, respectively, with 248 (68.9%) having uncontrolled BP. During visit two, 202 (56.1%) participants had uncontrolled BP, and their means SBP (mean SD) and DBP (mean SD) were 143.43±12.29 and 92.20±44.55, respectively, and there were 192 (53.3%) participants during visit three with means SBP (mean SD) and DBP (mean SD) 141.41±11.04 and 88.72±7.53, respectively. From patients’ self-response to the 9-item HB-MAS, the prevalence of antihypertensive medication adherence was 172 (47.8%).

**Table 2.**
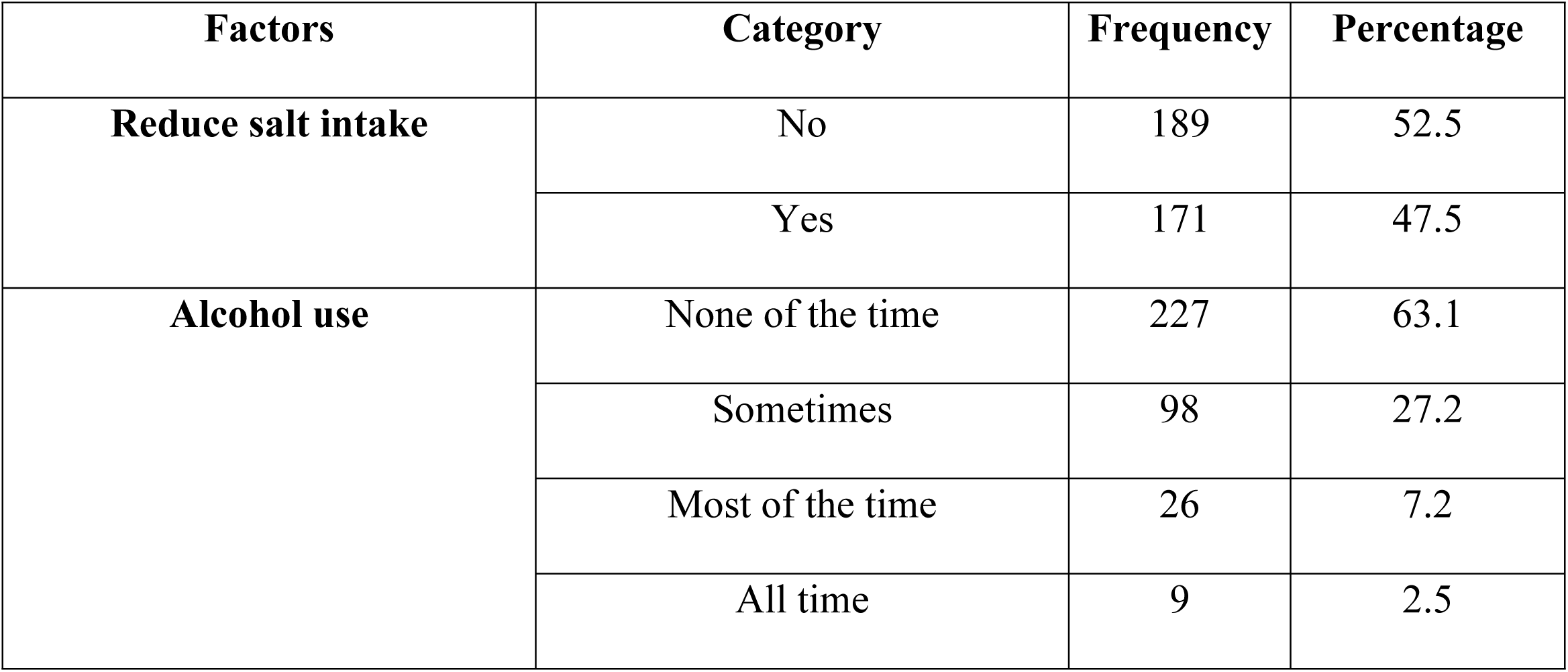
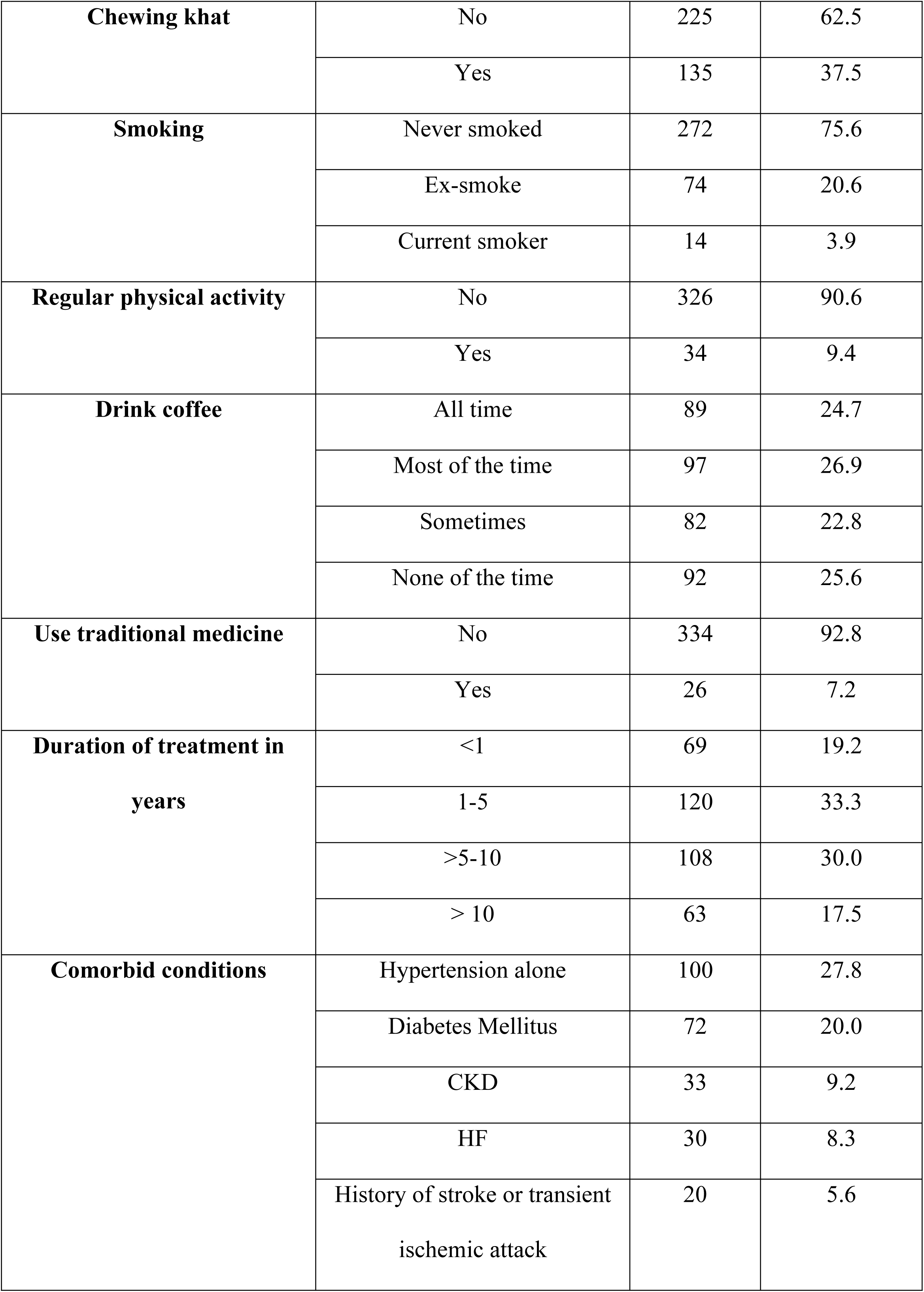
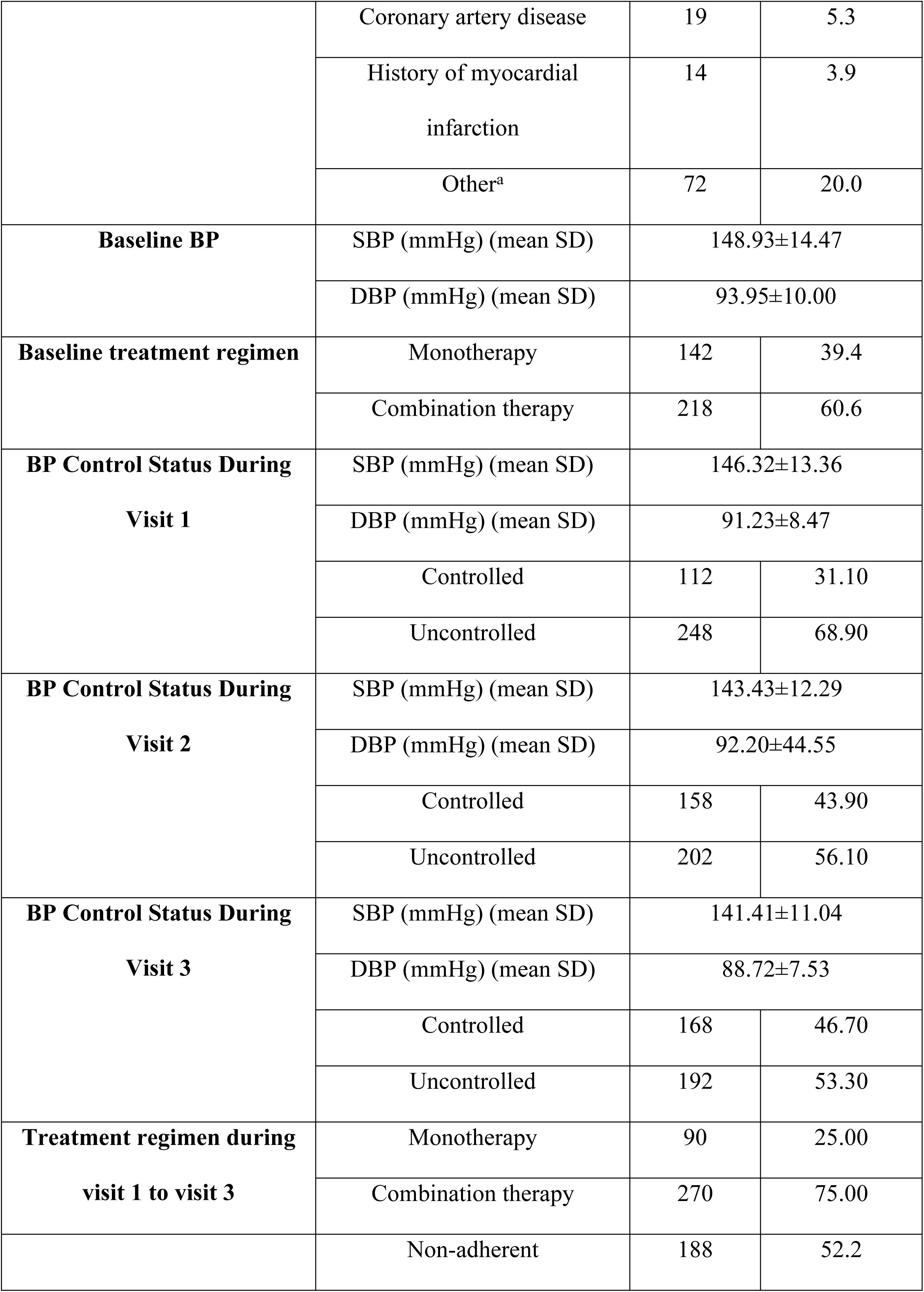
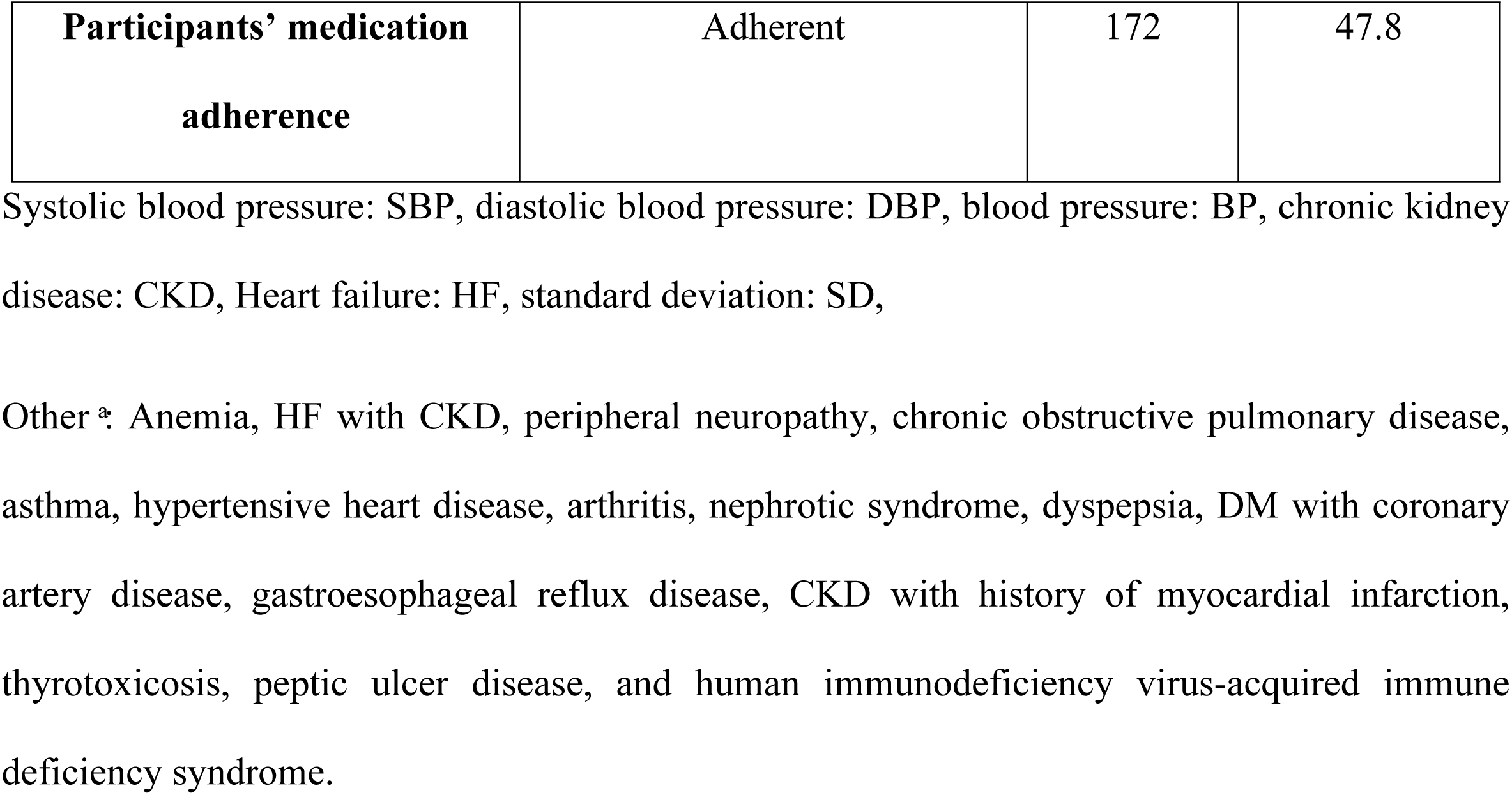
Lifestyle-related factors, clinical characteristics, and medication adherence of hypertensive participants at JMC from September 22 to December 22, 2022.

### Therapeutic inertia

Table 3 below shows the participants’ three-month mean (SD) TI score of 0.32 ± 0.51. Of the 360 participants, more than half, 205 (56.9%), had higher TI Scores. Antihypertensive treatment changes were not made in 104 (28.9%) participants with uncontrolled BP throughout their follow-up.

**Table 3.** TI Score and treatment changes were made among hypertensive participants at JMC from September 22 to December 22, 2022.

| Variables | Category | Frequency | Percentage |
| --- | --- | --- | --- |
| Mean TI Score | Mean ± SD | $0.32 \pm 0.51$ | |
|  | Lower | 155 | 43.1 |
|  | Higher | 205 | 56.9 |
| **Treatment changes made throughout the Follow-up** | Increased or added medication class whenever the BP is raised | 256 | 71.1 |
|  | No increased or added medication class whenever BP is raised | 104 | 28.9 |

Therapeutic inertia: TI, blood pressure: BP, standard deviation: SD.

As shown in Table 4, out of the total participants, 235 (65.3%) participants had a higher TI Score during visit one. During the remaining visits, participants had a higher TI Score (48.6%). Treatment changes were not made during visit one in nearly two-thirds (65.0%) of participants with uncontrolled BP. During the remainder of the visits, almost half of the participants had no increased or added medication class when their BP was raised. In 12 (3.3%), 18 (5.0%), and 7 (1.9%) participants in whom treatment change was made, BP was still uncontrolled during visits 1, 2, and 3, respectively. Among the factors related to TI, medication adherence concerns accounted for the majority, followed by medication complexity throughout the follow-up. Other factors included poor lifestyle adherence, traditional drug use, cost of medication, and poor communication with a clinician.

**Table 4.** TI Score, treatment changes made, and factors related to TI among hypertensive participants during visits 1, 2, and 3 at JMC from September 22 to December 22, 2022.

| Variables | Category | During Visit 1 |  | During Visit 2 |  | During Visit 3 |  |
| --- | --- | --- | --- | --- | --- | --- | --- |
|  |  | Frequency | Percentage | Frequency | Percentage | Frequency | Percentage |
|  | Lower | 125 | 34.7 | 185 | 51.4 | 185 | 51.4 |
|  | Higher | 235 | 65.3 | 175 | 48.6 | 175 | 48.6 |
| **TI Score**<br>**during**<br>**each visit** |  |  |  |  |  |  |  |
| **Treatment**<br>**changes**<br>**made**<br>**during**<br>**each visit** | Increased or added medication class whenever the BP is raised | 111 | 30.8 | 65 | 18.1 | 58 | 16.1 |
|  | No increased or added medication class whenever the BP is raised | 234 | 65.0 | 174 | 48.3 | 173 | 48.1 |
|  | BP was already controlled | 3 | 0.8 | 103 | 28.6 | 122 | 33.9 |
|  | Increased or added medication class whenever the BP was raised but remained uncontrolled | 12 | 3.3 | 18 | 5.0 | 7 | 1.9 |
| **Factors**<br>**related to**<br>**TI** | Medication adherence concern | 133 | 36.9 | 101 | 28.1 | 102 | 28.3 |
|  | Medication complexity concern | 36 | 10.0 | 31 | 8.6 | 39 | 10.8 |
|  | The participant prefers no treatment or changes in treatment | 18 | 5.0 | 12 | 3.3 | 12 | 3.3 |
|  | Unsure about true BP | 14 | 3.9 | 9 | 2.5 | 7 | 1.9 |
|  | Competing priorities | 8 | 2.2 | 4 | 1.1 | 5 | 1.4 |
|  | Other* | 27 | 7.5 | 17 | 4.7 | 8 | 2.2 |

Blood pressure: BP
Other\*: Poor lifestyle adherence, traditional drug use, unaffordability of medication, and poor communication with clinicians.

### Prescription patterns of antihypertensive medications

Of the participants, 90 (25.0%) were on monotherapy, and 270 (75.0%) were on combination therapy during the study period. Enalapril was the most frequently prescribed monotherapy drug, 166 (15.4), throughout the study. Hydrochlorothiazide (HTC) 77 (7.1%) and amlodipine 65 (6.0%) were monotherapy drugs other than Enalapril that were prescribed nearly equally in this study, as shown in Table 5 below.

**Table 5.** Frequency of monotherapy drugs prescribed during each visit among hypertensive participants at JMC from September 22 to December 22, 2022.

| Name of drug | Frequency | Percentage |
| --- | --- | --- |
| Enalapril | 166 | 15.4 |
| **Hydrochlorothiazide** | 77 | 7.1 |
| **Amlodipine** | 65 | 6.0 |
| **Losartan** | 6 | 0.6 |
| **Metoprolol** | 2 | 0.2 |

Hydrochlorothiazide: HTC

The mainly prescribed two-drug combination therapy was Enalapril + HCT 45 (12.5%), 55 (15.3%), and 59 (16.4%) during visit one, two, and three, respectively, followed by Amlodipine + HCT 43 (12.0%), 48 (13.4%), and 51(14.2%), during visit one, two, and three, and Amlodipine + Enalapril 44 (12.2%), 47 (13.0%), and 46 (12.8%), during visit one, two, and three, respectively, as shown in Table 6 below.

**Table 6.** Frequency of two drug combinations prescribed during each visit among hypertensive participants at JMC, 2022.

| **Drugs** | **Visit 1** |  | **Visit 2** |  | **Visit 3** |  |
| --- | --- | --- | --- | --- | --- | --- |
|  | **Frequency** | **Percentage** | **Frequency** | **Percentage** | **Frequency** | **Percentage** |
| **Amlodipine + HCT** | 43 | 12.0 | 48 | 13.4 | 51 | 14.2 |
| **Amlodipine + Furosemide +**<br>**Spironolactone** | 1 | 0.3 | 1 | 0.3 | 1 | 0.3 |
| **Amlodipine + Enalapril** | 44 | 12.2 | 47 | 13.0 | 46 | 12.8 |
| **Amlodipine + Metoprolol** | 3 | 0.8 | 3 | 0.8 | 3 | 0.8 |
| **Amlodipine + Propranolol** | 2 | 0.6 | 1 | 0.3 | 3 | 0.8 |
| **HCT + Enalapril** | 45 | 12.5 | 55 | 15.3 | 59 | 16.4 |
| **Furosemide + Enalapril** | 3 | 0.8 | 3 | 0.8 | 2 | 0.6 |
| **Losartan + Metoprolol** | 1 | 0.3 | 1 | 0.3 | 1 | 0.3 |
| **HCT + Losartan** | 3 | 0.8 | 5 | 1.4 | 5 | 1.4 |
| **Furosemide +**<br>**Spironolactone + Metoprolol** | 1 | 0.3 | 1 | 0.3 | 1 | 0.3 |
| **Furosemide + Atenolol** | 1 | 0.3 | 1 | 0.3 | 1 | 0.3 |
| **Furosemide + Propranolol** | 2 | 0.6 | 1 | 0.3 | 1 | 0.3 |
| **Enalapril + Atenolol** | 1 | 0.3 | 1 | 0.3 | 1 | 0.3 |
| **Enalapril + Metoprolol** | 18 | 5.0 | 18 | 5.0 | 18 | 5.0 |
| **Enalapril + Propranolol** | 1 | 0.3 | 1 | 0.3 | 1 | 0.3 |

Hydrochlorothiazide: HTC

As shown in Table 7, among the three-drug combination therapies, Amlodipine + Enalapril + HCT (19 (5.3%), 18 (5.0%), and 18 (5.0%) during visits one, two, and three, respectively) was the most commonly prescribed three-drug combination therapy, followed by Amlodipine + Losartan + HCT (5 (1.4%), 7 (2.0%), and 9 (2.2%) during visits one, two, and three, respectively). Of the four-drug combination therapies, Enalapril + Amlodipine + Metoprolol + HCT (7 (1.9%) during all three visits) was the most frequently prescribed in this study.

**Table 7.**
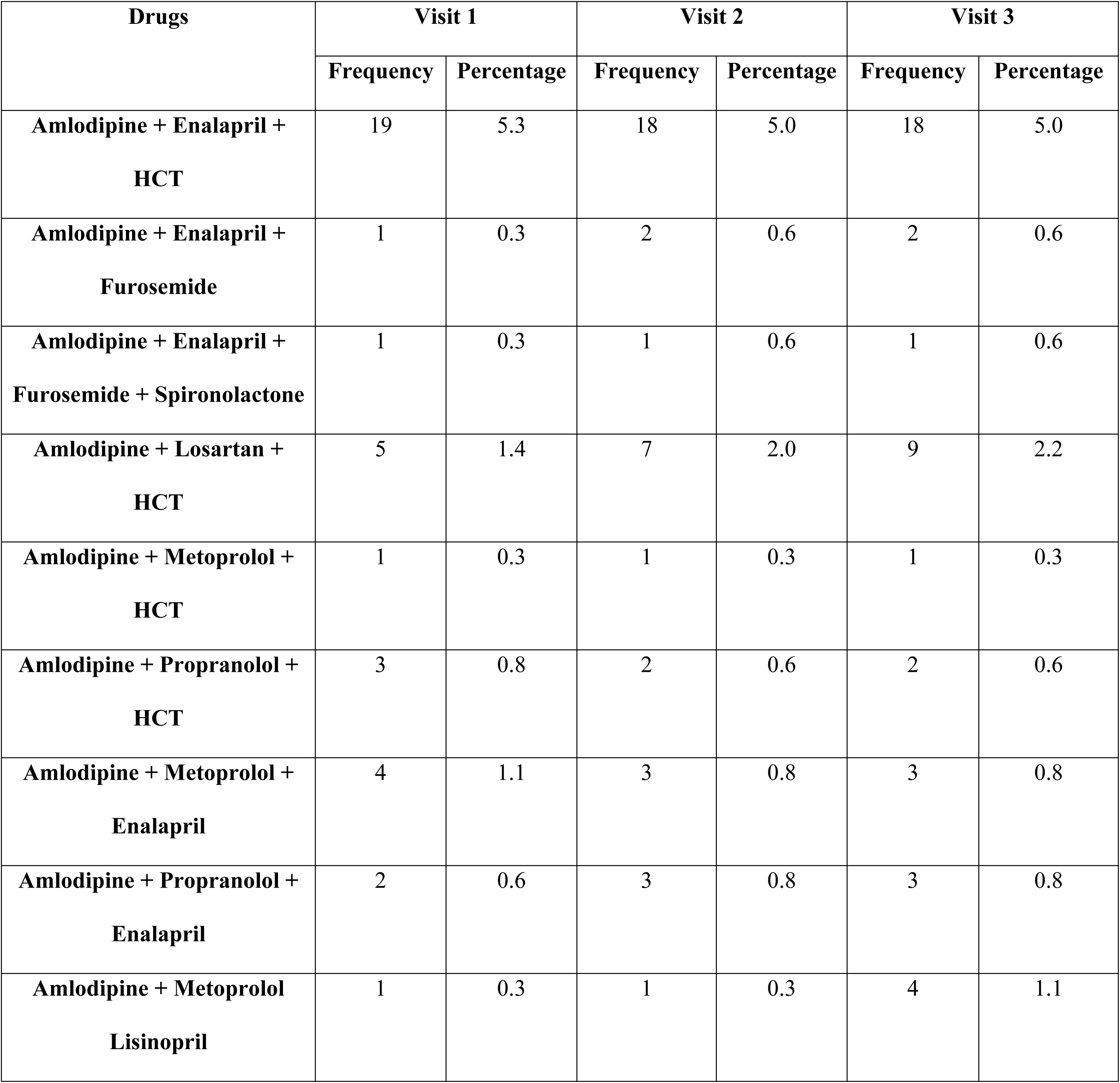
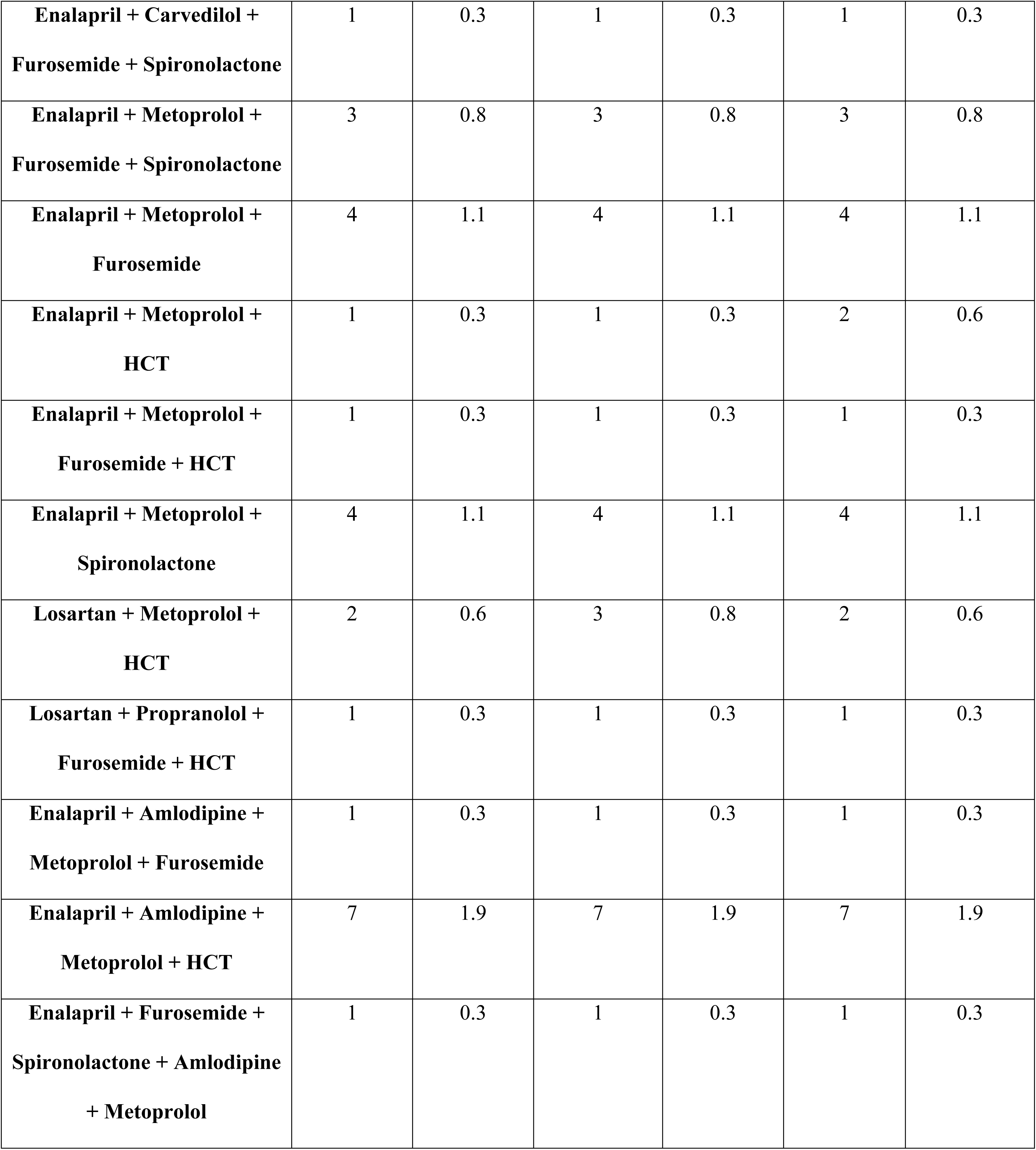
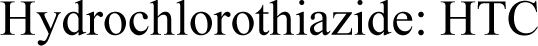
Frequency of ≥3 drug combinations prescribed during each visit among hypertensive participants at JMC from 22 September to 22 December 2022.

| Drugs | Visit 1 |  | Visit 2 |  | Visit 3 |  |
| --- | --- | --- | --- | --- | --- | --- |
|  | Frequency | Percentage | Frequency | Percentage | Frequency | Percentage |
| **Amlodipine + Enalapril +**<br>**HCT** | 19 | 5.3 | 18 | 5.0 | 18 | 5.0 |
| **Amlodipine + Enalapril +**<br>**Furosemide** | 1 | 0.3 | 2 | 0.6 | 2 | 0.6 |
| **Amlodipine + Enalapril +**<br>**Furosemide + Spironolactone** | 1 | 0.3 | 1 | 0.6 | 1 | 0.6 |
| **Amlodipine + Losartan +**<br>**HCT** | 5 | 1.4 | 7 | 2.0 | 9 | 2.2 |
| **Amlodipine + Metoprolol +**<br>**HCT** | 1 | 0.3 | 1 | 0.3 | 1 | 0.3 |
| **Amlodipine + Propranolol +**<br>**HCT** | 3 | 0.8 | 2 | 0.6 | 2 | 0.6 |
| **Amlodipine + Metoprolol +**<br>**Enalapril** | 4 | 1.1 | 3 | 0.8 | 3 | 0.8 |
| **Amlodipine + Propranolol +**<br>**Enalapril** | 2 | 0.6 | 3 | 0.8 | 3 | 0.8 |
| **Amlodipine + Metoprolol**<br>**Lisinopril** | 1 | 0.3 | 1 | 0.3 | 4 | 1.1 |
| **Enalapril + Carvedilol +**<br>**Furosemide + Spironolactone** | 1 | 0.3 | 1 | 0.3 | 1 | 0.3 |
| **Enalapril + Metoprolol +**<br>**Furosemide + Spironolactone** | 3 | 0.8 | 3 | 0.8 | 3 | 0.8 |
| **Enalapril + Metoprolol +**<br>**Furosemide** | 4 | 1.1 | 4 | 1.1 | 4 | 1.1 |
| **Enalapril + Metoprolol +**<br>**HCT** | 1 | 0.3 | 1 | 0.3 | 2 | 0.6 |
| **Enalapril + Metoprolol +**<br>**Furosemide + HCT** | 1 | 0.3 | 1 | 0.3 | 1 | 0.3 |
| **Enalapril + Metoprolol +**<br>**Spironolactone** | 4 | 1.1 | 4 | 1.1 | 4 | 1.1 |
| **Losartan + Metoprolol +**<br>**HCT** | 2 | 0.6 | 3 | 0.8 | 2 | 0.6 |
| **Losartan + Propranolol +**<br>**Furosemide + HCT** | 1 | 0.3 | 1 | 0.3 | 1 | 0.3 |
| **Enalapril + Amlodipine +**<br>**Metoprolol + Furosemide** | 1 | 0.3 | 1 | 0.3 | 1 | 0.3 |
| **Enalapril + Amlodipine +**<br>**Metoprolol + HCT** | 7 | 1.9 | 7 | 1.9 | 7 | 1.9 |
| **Enalapril + Furosemide +**<br>**Spironolactone + Amlodipine**<br>**+ Metoprolol** | 1 | 0.3 | 1 | 0.3 | 1 | 0.3 |

Hydrochlorothiazide: HTC

### BP control levels among hypertensive participants

Based on the 8^th^ JNC guideline recommendations, BP control was achieved in 133 (36.9%) hypertensive participants. Of the total study participants, 248 (68.9%), 202 (56.1%), and 192 (53.3%) during visit one, visit two, and visit three had uncontrolled BP, respectively, as shown in Fig. 2 below.

**Fig 2.**
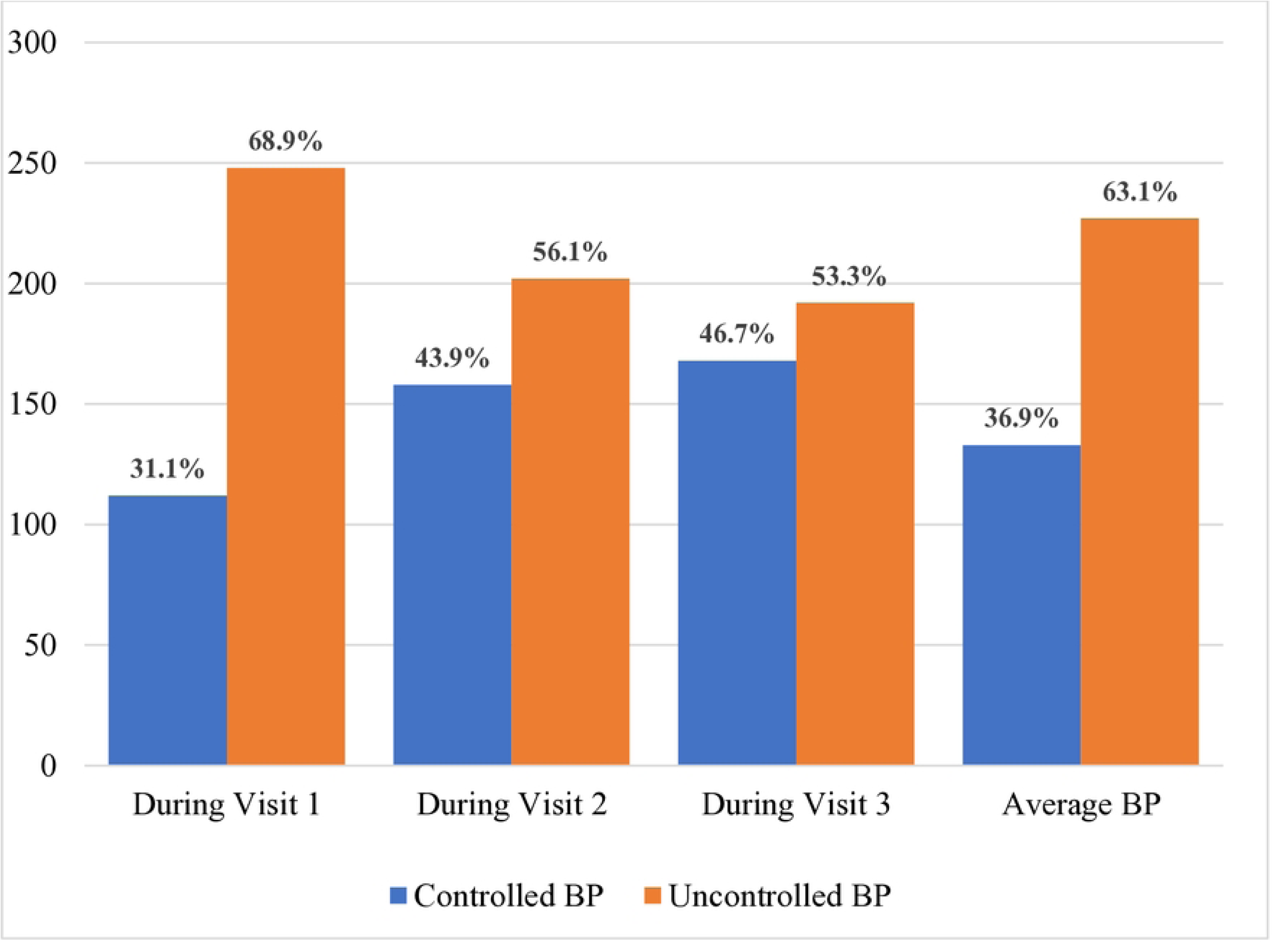
Blood pressure control rates among hypertensive participants at JMC from 22 September to 22 December 2022. The blue bars indicate patients with controlled BP, while the orange bars show patients with uncontrolled BP.

As shown in Table 8 below, the overall mean SD of SBP was 143.73 ± 11.30 mm Hg, while the DBP was 90 ± 6.87 mm Hg.

**Table 8.** Mean BP control level among hypertensive participants at JMC from 22 September to 22 December 2022.

| Description | At Visit 1 | At Visit 2 | At Visit 3 | Average BP |
| --- | --- | --- | --- | --- |
| SBP (mmHg) (mean SD) | $146.31 \pm 13.32$ | $143.37 \pm 12.29$ | $141.45 \pm 11.04$ | $143.73 \pm 11.30$ |
| DBP (mmHg) (mean SD) | $91.26 \pm 8.514$ | $89.93 \pm 8.34$ | $88.89 \pm 8.38$ | $90 \pm 6.87$ |

Systolic blood pressure: SBP, diastolic blood pressure: DBP, standard deviation: SD

### Determinants of BP control among hypertensive participants

Variables with a P-value below 0.25 in bivariate logistic regression analysis were age, residency, salt use, khat chewing, coffee drinking, traditional drug use, duration of treatment, baseline blood pressure status, baseline antihypertensive regimens, BP status during each visit, average SBP and average DBP, treatment regimen, TI Score during each visit, mean TI Score, and medication adherence status. Hence, these variables were incorporated into the multivariate logistic regression model for further analysis. However, following multivariate regression analysis, not salt reducing, medication nonadherence, and TI Score during each visit, and the mean TI Score were identified as independent variables significantly associated with uncontrolled BP, as shown in Table 9 below. Accordingly, Participants who were not reducing salt intake had about 6 times (AOR = 5.96, 95% CI = 1.86-19.12, *P* = 0.003) more odds of uncontrolled BP compared with participants reducing salt in their diet, and medication-non-adherent participants had 4.37 times greater odds of having uncontrolled BP than adherents (AOR = 4.37, 95% CI = 1.43-13.30, *P* = 0.009). TI Score at visits 1, 2, and 3 were 7.86 times (AOR = 7.86, 95% CI = 2.37-26.05, *P* = 0.001), 5.85 times (AOR = 5.85, 95% CI = 1.44-23.72, *P* = 0.013), and 3.98 times (AOR = 3.98, 95% CI = 1.06-14.92, *P* = 0.041), respectively, and significantly associated with uncontrolled BP. Additionally, compared to those with a lower TI Score, those with a greater TI Score had a reduced likelihood of having controlled BP (AOR = 0.11, 95% CI = 0.02-0.63, *P* = 0.013).

**Table 9.**
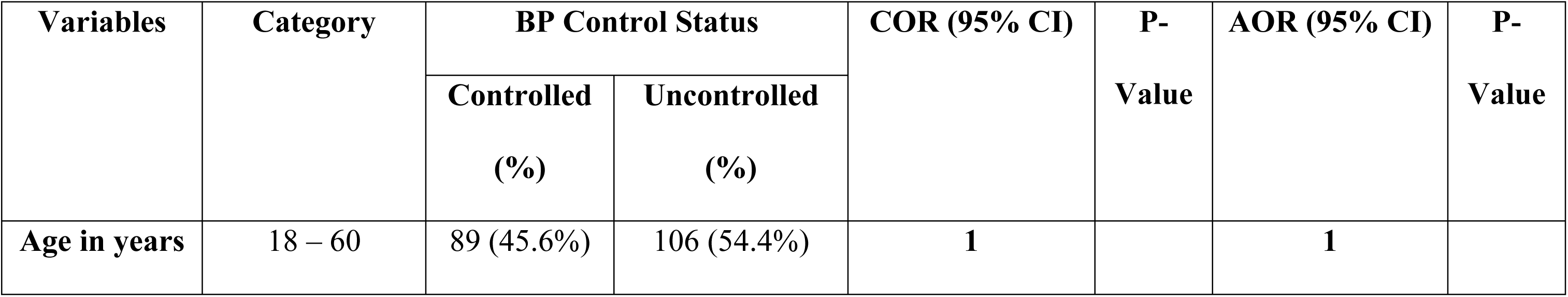
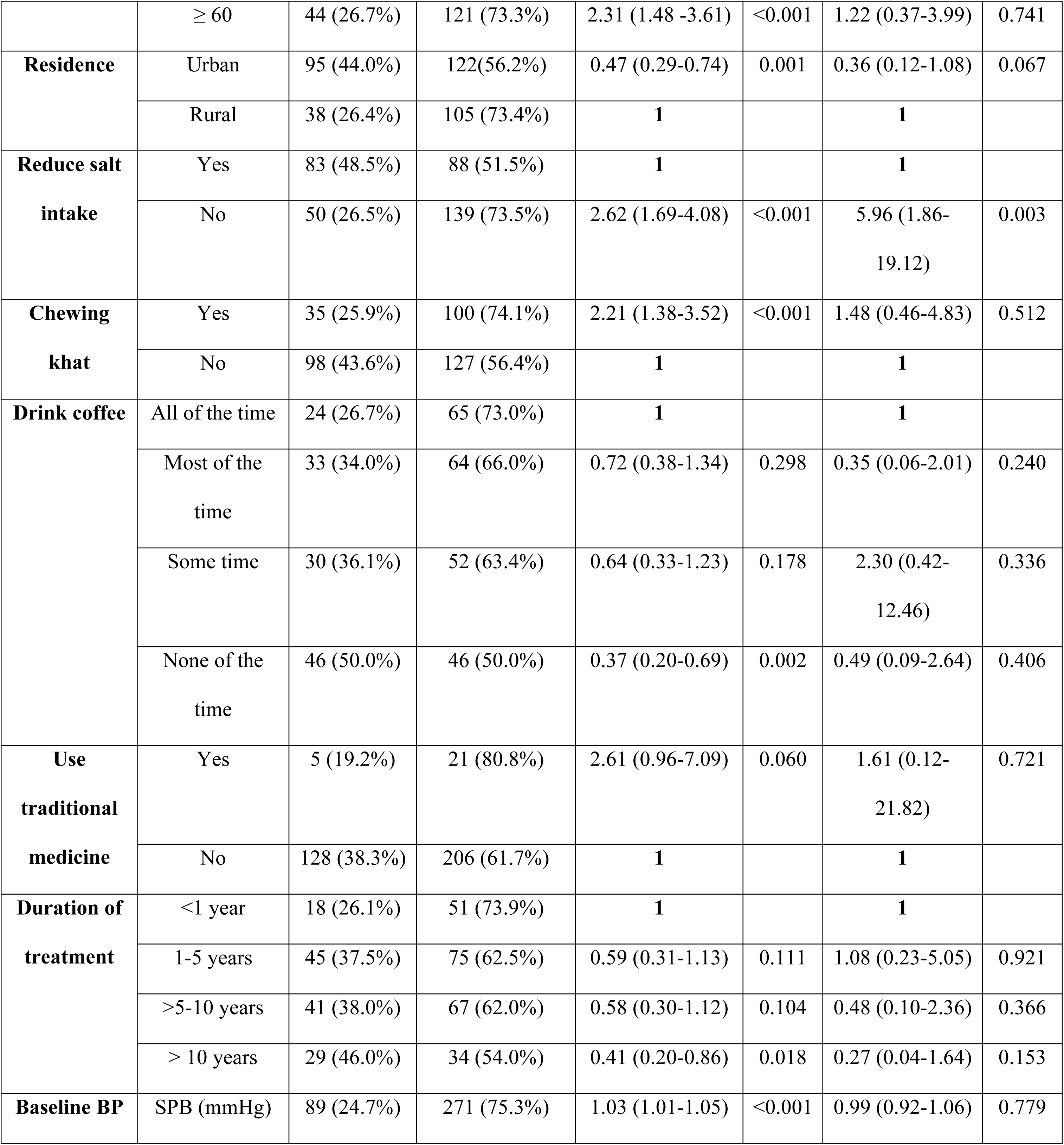
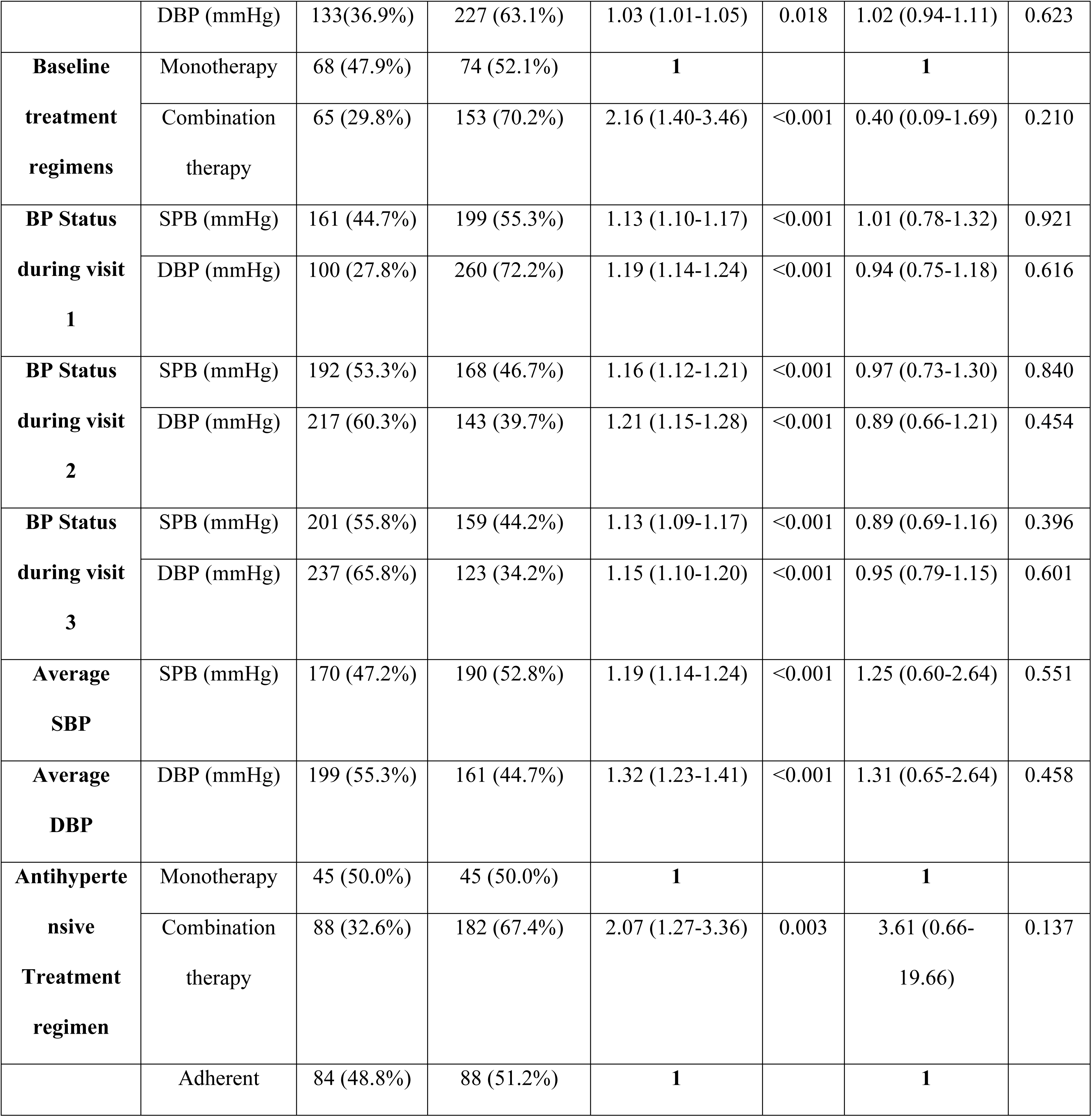
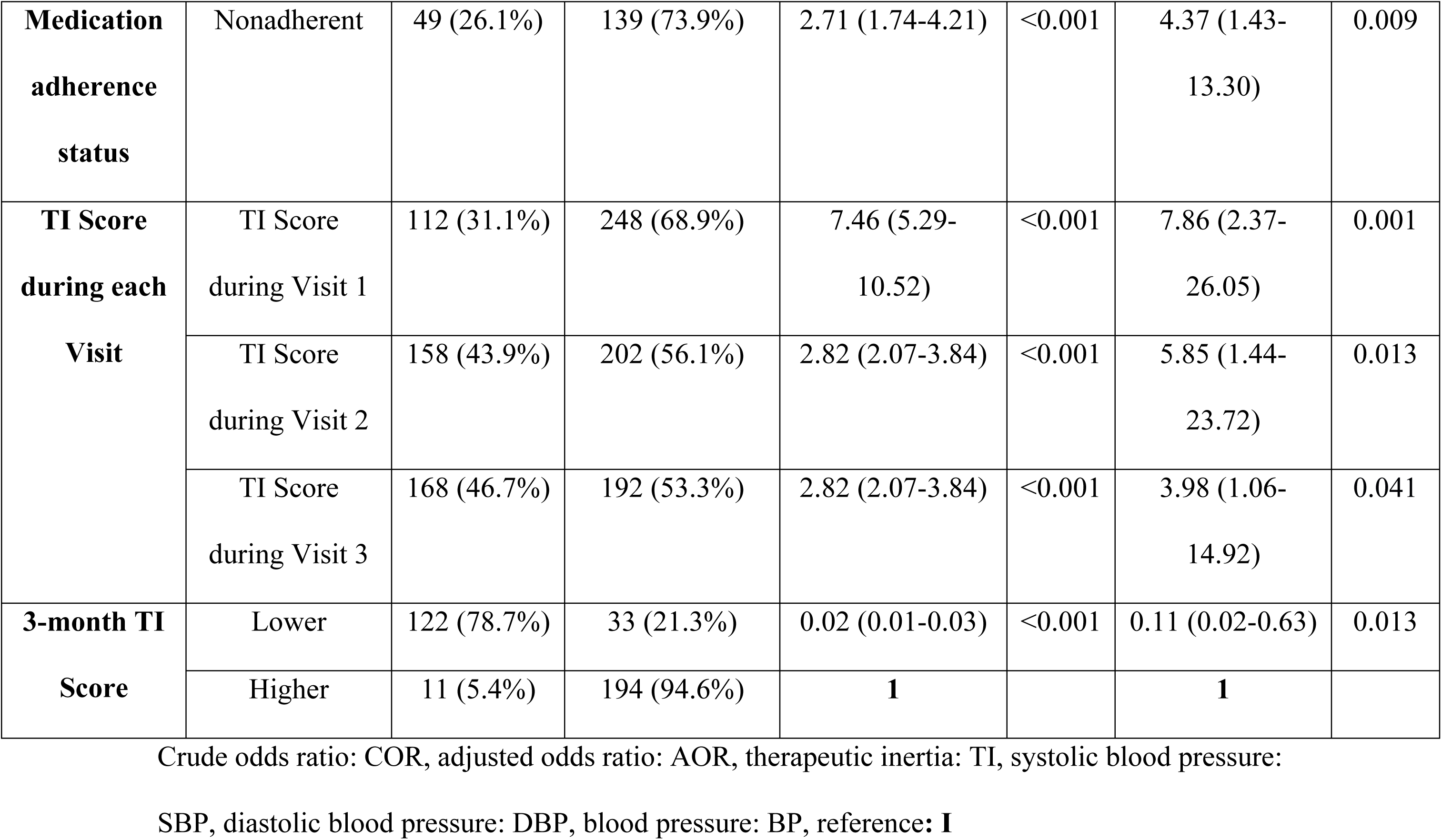
Binary logistic and multiple regression analyses to identify factors associated with uncontrolled BP among hypertensive participants at JMC from September 22 to December 22, 2022.

## Discussion

Hypertension remains a significant public health issue. The rate of BP control among hypertension patients is poor, including in Ethiopia. The current study examined the burden of TI and the BP control status among adult hypertension patients attending follow-up at the Ambulatory Cardiac Clinic of JMC. This study confirms that uncontrolled hypertension is a serious public health concern, as less than half (36.9%) of hypertensive patients had controlled BP while on routine follow-ups at the hospital, highlighting that uncontrolled hypertension is a significant health problem.

The findings of this study were the same as the results of the study conducted in six public hospitals in Addis Ababa and Tigray regional state, Ethiopia (37.0%) (42). This could be related to the socioeconomic similarity of the participants. The findings were also almost consistent with studies in different areas of Ethiopia and Tanzania (42.8%, 42.9%, and 43.2%) (31, 33, 43). This could be related to lifestyle factors similar to those in this study. In contrast, the rate of control of BP found in this study was smaller than that found in Sudan (64.0%), Southwest Ethiopia (50.3%), eastern Sudan (45.3%), and Kedah (48.3%) (32, 44–46). The higher BP control rate can be related to improved medical and medication management for participants, improved education and knowledge of participants towards hypertension treatment, and implementing different recommendations as a standard in these countries. However, the rate of blood pressure control was better compared to studies done in Addis Ababa (31%) and Nigeria (27.9%) (47, 48). The disparity can be attributed to discrepancies in inclusion criteria and the sites of studies. Moreover, two studies used a cross-sectional study design, which could have played a role in this variation. The findings were also larger than those of the studies conducted in Morocco and Addis Ababa, Ethiopia (27% and 30.1%) (49, 50). This gap could be attributed to changes in the study design, degree of urbanization, lifestyle habits, and eating habits.

There was a strong association between TI and uncontrolled BP, as shown by this study. This study found that the TI Score during visits 1, 2, and 3 was 7.86, 5.85, and 3.98 times more likely to be associated with uncontrolled BP. In addition, participants with a higher TI Score were 89% less likely to have controlled BP than participants with a lower TI Score. The findings of this study are comparable to the studies from Morocco, Sub-Saharan African countries, the United States of America, China, and Spain (21, 22, 36, 50, 51). The possible reasons may be an overemphasis on the effect of the recommended treatments, the clinician’s doubts about the adherence, preventing the decision to intensify the treatment, or a lack of clinician training to achieve the therapeutic desired outcome (52). In addition, the impact of the economic and social status of the patient with hypertension on the medical choices, the lack of adequate antihypertensive medications at public health facilities, and failure to adhere to the recommended treatments for managing hypertensive patients may also be possible reasons. This study additionally showed that medication non-adherent participants had more than four times the likelihood of having uncontrolled BP than adherents. This finding matched studies conducted in Ethiopia, Ghana, Morocco, and Sudan (33, 38, 45, 50, 53, 54). Reasons for nonadherence are multifactorial and are often contributed to by a blend of patient-related, physician-related, and health system-related factors. This study also shows a strong association between adding salt to food and uncontrolled BP. Accordingly, compared to participants who reduced their salt intake, participants who did not do so had a roughly six times higher chance of having uncontrolled BP. This result aligned with studies undertaken in different parts of Ethiopia (31, 32, 53). This may be because high salt intake induces fluid retention, raising cardiac strain and high BP.

### Strengths and limitations of the study

This study has notable strengths. First, it involved a relatively large sample size and was completed over three months, unlike previous studies at JMC. Second, it examined the burden of TI and aimed to determine the rates of BP control and TI. Third, it sought to identify factors associated with poor BP management. Fourth, the results highlight significant issues regarding the impact of TI on inadequate hypertension control. TI is clearly vital for understanding and addressing poor hypertension management. Consequently, these findings can serve as a foundation for future research. However, there are some limitations. Despite its strengths, this study does not evaluate health professionals’ knowledge about hypertension management, clinicians’ adherence to guidelines, participants’ knowledge, attitudes, and perceptions of hypertension treatment, nor does it consider concomitant medications or adverse effects of drugs. Additionally, since the study was conducted at a single facility, the results might not apply to the broader healthcare setting across Ethiopia.

### Conclusion

The rate of controlled BP was low. Not reducing salt, medication non-adherence, and TI were independent predictors of BP control. Therefore, clinicians should follow guidelines to reduce TI and improve treatment and care for hypertensive patients. Clinicians should also educate patients about salt intake and effective medication adherence. Finally, ongoing focus and research are needed to explore the potential impact of TI in managing HTN.

## Data Availability

All relevant data are within the manuscript and its Supporting Information files.

## Acknowledgments

We thank the participants of the study.

## Supporting information

**S1 File.** Data collection tool. Used to collect informations from patient and clinicians.

**S1 Dataset.** Data.

